# Epigenetic Age Monitoring in Professional Soccer Players for Tracking Recovery and the Effects of Strenuous Exercise

**DOI:** 10.1101/2024.11.28.24317877

**Authors:** Robert T Brooke, Thomas Kocher, Roland Zauner, Juozas Gordevicius, Milda Milčiūtė, Marc Nowakowski, Christian Haser, Thomas Blobel, Johanna Sieland, Daniel Langhoff, Winfried Banzer, Steve Horvath, Florian Pfab

**Affiliations:** Epigenetic Clock Development Foundation, Torrance, CA, USA; EB House Austria, Research Program for Molecular Therapy of Genodermatoses, Department of Dermatology and Allergology, University Hospital of the Paracelsus Medical University Salzburg, 5020 Salzburg, Austria; DNAthlete Austria GmbH, 5020 Salzburg, Austria; DNAthlete AG, 9494 Schaan, Lichtenstein; Eintracht Frankfurt Fußball AG, 60528 Frankfurt am Main, Germany; Eurofins Genomics Europe, Brendstrupgaardsvej 23, DK-8200 Aarhus N; Division of Preventive and Sports Medicine, Institute of Occupational, Social and Environmental Medicine, Goethe University Frankfurt, Frankfurt/Main, Germany; Altos Labs, Cambridge, UK; Technische Universitaet Munich, Munich, Germany; Brighton & Hove Albion Football Club, Brigthon, UK

**Keywords:** FitAge, GrimAge, Soccer, Sports Injury, Exercise, C-Reactive Protein

## Abstract

Elite sports have become increasingly professionalized and personalized, with soccer players facing a high number of games per season. This trend presents significant challenges in optimizing training for peak performance and requires rigorous monitoring of athletes to prevent overload and reduce injury risks. The emerging field of epigenetic clocks offers promising new pathways for developing useful biomarkers that enhance training management. This study investigates the effects of intense physical activity on epigenetic age markers in professional soccer players. We analyzed DNA methylation data from saliva samples collected before and after physical activity. Vigorous physical activity was found to rejuvenate epigenetic clocks, with significant decreases in DNAmGrimAge2 and DNAmFitAge observed immediately after games. Among player subgroups, midfielders exhibited the most substantial epigenetic rejuvenation effect following games. Additionally, the study suggests a potential link between DNA methylation patterns and injury occurrence. Overall, our study suggests that DNA methylation based biomarkers may have applications in monitoring athlete performance and managing physical stress.

## 1 Introduction

Elite sports have experienced an enormous surge in professionalization and personalization of competition, training, prevention and recovery management. The demands on athletes have increased owing to the evolving nature of sports and tight competition schedules, with elite soccer players playing up to 75 games per season. Beside tight time schedules and extensive travel, soccer has undergone a significant evolution in terms of the dynamic nature of the game, with increased running distances, number of runs, number of sprints and high-speed actions (Barnes et al. 2014, Wallace et al. 2014, Haller et al. 2023). In recent decades, a variety of performance parameters have been established and considered to support the decision-making processes for coaches and physicians, with regard to acute load and recovery management. External load can be expressed and monitored through time-motion analysis, tracking devices, or power parameters, like covered distance or peak power output. In addition, team physicians often rely on practical, scientifically well-researched, and rapidly measurable biomarkers such as creatine kinase (CK) or lactate (Haller et al. 2023). High levels of CK and/or high-sensitivity interleukin (IL)-6 levels can result from inflammatory processes due to excessive exercise load (Romagnoli et al. 2016, Thorpe et al. 2012). These biomarkers can be used to assess the acute internal load by assessing tissue- or organ-specific fatigue, stress, damage and/or recovery processes (Haller et al. 2023).

Effectively monitoring and managing training load is essential not only for optimizing athletic performance but also for preventing overtraining and reducing the risk of injuries. Sports-related injuries represent a significant healthcare burden and can lead to considerable psychological and motivational setbacks for athletes. Additionally, in professional sports, injuries have substantial economic repercussions (Ryan et al. 2019; Trentacosta et al. 2020). The predominant etiology of sports-related injuries is overuse injuries, resulting from repetitive stress and micro-traumas, sometimes culminating in severe traumatic injuries (Tarnowski et al. 2022). Genetic information has been shown to be associated with performance-related effects of sports training and predisposition to injuries (Guilherme et al. 2014, Varillas-Delgado et al. 2022, Ginevicien et al. 2022, Pfab et al. 2023).

This study examines whether epigenetic markers, specifically DNA methylation levels, can be used to develop indicators related to injury risk. Experimental evidence suggests that exercise acts as a significant stressor, driving various physiological adaptations in the body, including changes in epigenetic mechanisms. For example, research by Denham et al. demonstrated alterations in DNA methylation patterns in skeletal muscle following acute exercise, indicating a dynamic epigenetic response to exercise-induced stress (Denham et al. 2014). A study by Rönn et al revealed exercise-induced changes in DNA methylation in adipose tissue, particularly in genes related to metabolism and inflammation (Rönn et al. 2013). Changes in methylation patterns can be interpreted biologically, for example, by using epigenetic clocks. Epigenetic clocks are DNAm-based prediction methods for estimating age or mortality risk (Horvath et al. 2013, Horvath et al. 2018, Lu et al. 2019). Second generation epigenetic clocks such as DNAm-based GrimAge (DNAmGrimAge) and GrimAge2 (DNAmGrimAge2) predict future morbidity and mortality risk (Lu et al. 2019, Li et al. 2020). While DNAmGrimAge was trained on blood samples and an older population, DNAmGrimAge2 is also applicable to younger individuals and saliva samples (Lu et al. 2022). DNAmGrimAge is a composite biomarker (weighted linear combination) of seven DNAm surrogates of plasma proteins, a DNAm-based estimator of smoking pack-years, age, and sex. The seven DNAm-based proteins comprise adrenomedullin (ADM), beta-2-microglobulin (B2M), cystatin C (Cystatin C), GDF-15, leptin (Leptin), plasminogen activator inhibitor 1 (PAI-1), and tissue inhibitor metalloproteinases 1 (TIMP- 1). Version 2 of GrimAge leverages two additional DNAm-based estimators of plasma proteins: CRP and hemoglobin A1C (logA1C) (Lu et al. 2022). Both versions of DNAmGrimAge perform well on predicting functional decline and onset of major age-related conditions reliably across large diverse populations, including heart disease, cancer onset, multi-modal measures of brain health, kidney disease, fatty liver, respiratory function, and more (Lu et al. 2019, Hillary et al. 2018, Hillary et al. 2020, McCrory et al. 2020). Another epigenetic clock, which potentially has relevance to athlete performance, integrates fitness parameters into DNAmGrimAge2, to construct DNAm-based FitAge (DNAmFitAge), a physical fitness age predictor. DNAmFitAge includes blood-based DNAm biomarkers for fitness parameters like gait speed, maximum handgrip strength, forced expiratory volume in one second, and maximal oxygen uptake (McGreevy et al. 2023). Studies have revealed a correlation between physical fitness and biological age, as measured by DNA methylation age (DNAmFitAge) (Jokai et al. 2023).

Physically fit individuals tend to have a younger DNAmFitAge, which is associated with improved age-related health outcomes. These individuals have a lower risk of mortality, coronary heart disease, and experience increased periods of disease-free status, indicating better overall health maintenance as they age (McGreevy et al. 2023).

To the best of our knowledge no studies have been conducted to investigate the short-term dynamics of epigenetic clocks in professional soccer players. Furthermore, no data is available to evaluate possible relationships between epigenetic stress markers and injury events. In this explorative study we aimed to investigate the potential of newly developed epigenetic clocks such as DNAmGrimAge2 and DNAmFitAge as biomarker to monitor and help to manage athlete performance and to prevent unwanted side effects, like musculoskeletal damages or injuries.

Here we generated DNA methylation levels from saliva samples collected from professional soccer team members during a season, including time points with high and low physical stress. Widely used epigenetic clocks were compared to established injury/inflammation markers like CK, IL-6 and C-reactive protein (CRP), and tested whether DNAm-based predictions thereof can be associated with injury occurrence.

## 2 Results

### 2.1 Vigorous physical activity leads to a rejuvenating effect on epigenetic age predictions

Saliva samples (n=201) were collected from 24 members of a first league soccer club at nine different time points over a period of 6 months during the professional soccer seasons of 2021/22 and 2022/23 (Table 1).

**Table 1:** Overview of study participants. . Age refers to the actual chronological age of participants at the beginning of the season.

| | N | | mean age ± sd<br>(years) |
| --- | --- | --- | --- |
|  | samples | subjects |  |
| all | 156 | 19 | 26.23 ± 4.41 |
| forwarder | 30 | 5 | 26.75 ± 2.40 |
| midfielder | 54 | 6 | 26.42 ± 5.89 |
| defender | 45 | 5 | 24.49 ± 2.11 |
| goalkeeper | 27 | 3 | 29.51 ± 5.79 |
| supporting staff | 45 | 5 | 42.61 ± 3.80 |

Time points for sample collection were chosen to cover different physical stress states of athletes, so that samples can be grouped according to medium (before match) and high activity (immediately after game) phases. The state “rested” represents the training & recovery phase after match days (Figure 1). Using the HumanMethylationEPIC v1.0 BeadChip (Illumina, San Diego, CA) over 800,000 CpG sites within genomic DNA isolated from saliva were analyzed for their methylation status (DNAm). Various recently developed epigenetic markers of aging, known as epigenetic clocks, were calculated from the obtained methylation data. These include DNAmGrimAge2, DNAmFitAge, and Skin & Blood Clock (Lu et al. 2022, McGreevy et al. 2023, Horvath et al. 2018). To investigate the influence of strenuous physical activity on epigenetic age, we compared age predictions derived from samples (n=156) obtained from athletes (n=19, mean age ± standard deviation (sd): 26.23 ± 4.41) divided into three groups representing different physical stress states based on the timing of collection (Figure 1). A control group was established using samples (n=45) from 5 supporting staff members (mean age ± sd: 42.61 ± 3.80). Compared to active players, the supporting staff members experienced the same environmental factors such as traveling, logistical challenges and emotional distress, but lower levels of acute physical stress. GrimAge2 and FitAge predictions were calibrated (cal.) to the actual age range of players, referred to as GrimAge2 cal. and FitAge cal., respectively.

**Figure 1:**
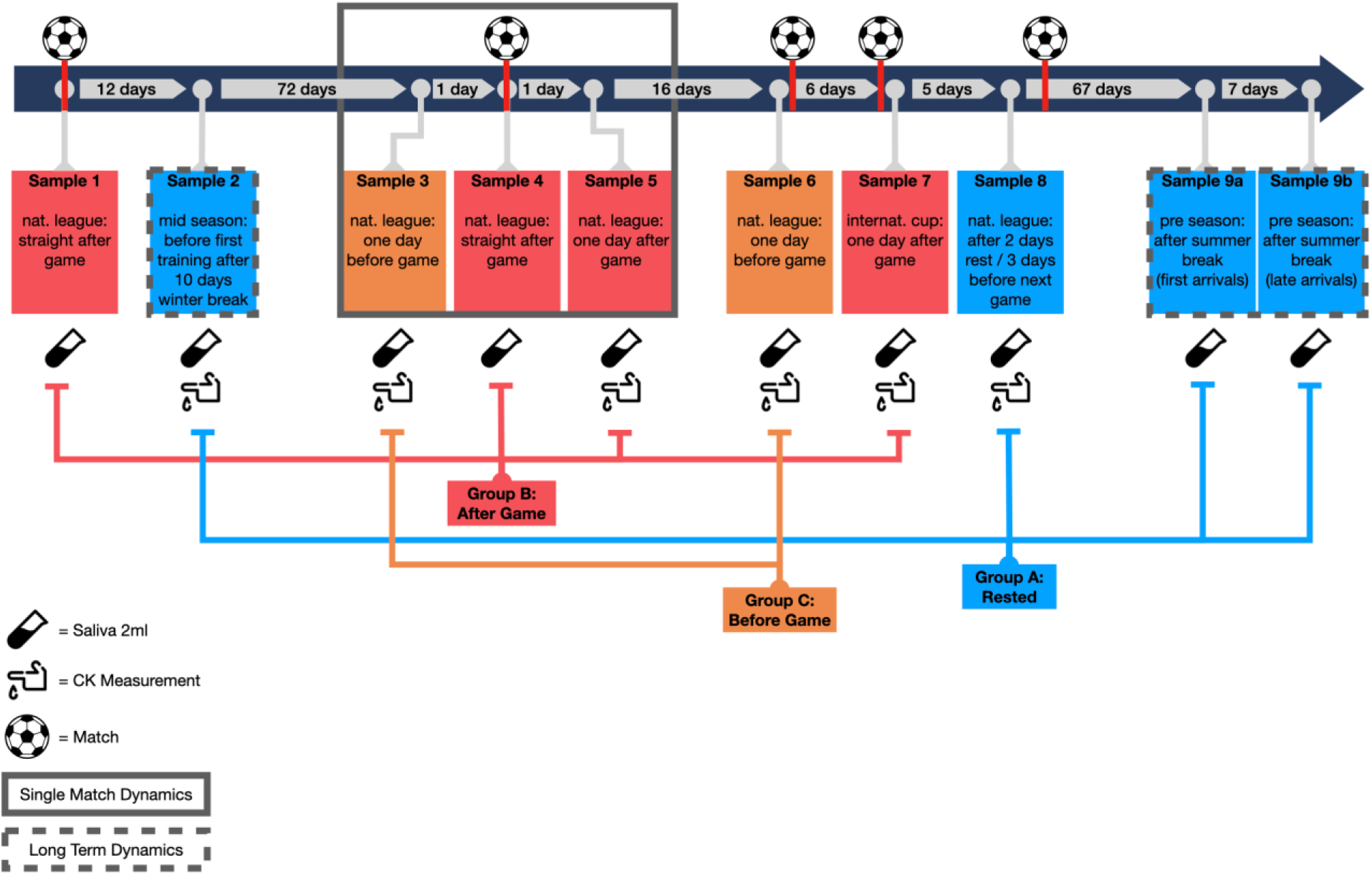
Sampling scheme and group assignment. Timeline indicates the individual measurement timepoints with respect to events and their intervals to each other. Events that involved strenuous physical exertion were denoted with a ball icon. Two different types of samples were collected from athletes as well as from supporting staff members serving as control group: saliva (indicated by saliva collection tube symbol) and blood samples for CK measurement (indicated by hand with blood drop symbol). DNA methylation data generated from saliva samples as well as CK measurement data was assigned to three groups: A - rested, B - straight or one day after match and C - one day before match. Sample 9 was split into a and b, as it was taken after the summer break and the players involved returned from the break at two different times. In addition to examining short-term effects after an intense game event (sample 3, 4 and 5; highlighted with a gray rectangle as “Single Match Dynamics”), data from sample 9 (end of season) was compared to data from sample 2 (start of season) to assess changes over a longer time frame (depicted with a dashed box as “Long Term Dynamics”).

Players’ DNAm patterns showed significant changes when comparing samples collected before and after the game throughout the season (Figure 2A,B). These changes led to a considerable decrease in biological age predictors: DNAmGrimAge2 calibrated (cal.) decreasing by 32% (p = 6.13e-05), and DNAmFitAge calibrated (cal.) by 18% (p = 0.00016).

**Figure 2:**
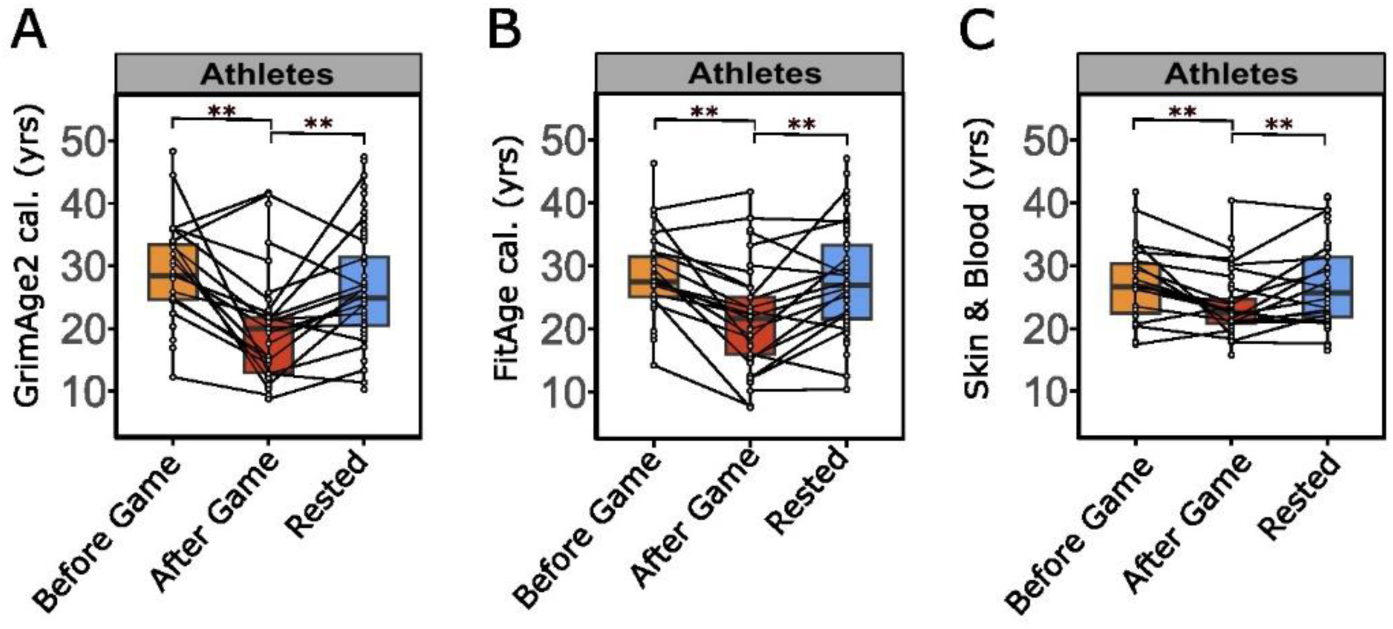
Intense physical activity causes rapid changes in biological age predictors. (A-C) Epigenetic profiles (DNAm) of saliva samples collected from athletes during medium to low physical stress states (yellow: before game, blue: rested) or immediately after intensive physical activity (red: after game) were used to estimate biological/chronological age in years (yrs), including **(A)** GrimAge2 cal. (before vs. after game p = 6.13e- 05; after game vs. rested p = 0.00281), **(B)** FitAge cal. (before vs. after game p = 0.00016; after game to rested p = 0.000476) and **(C)** chronological age predictor Skin & Blood Clock (before vs. after game p = 0.000307; after game vs. rested p = 3.19e-07). **(A-C)** Each dot represents predicted age in years (yrs) for a specific time point and player. Significance levels are indicated by * (p <= 0.05), ** (p <= 0.01, *** (p <= 0.001)) and NS. (p > 0.05). Statistical significance in predicted age differences was evaluated using a linear mixed effect model with chronological age, timepoint (before, after game or rested) and batch (sample processing batches) as fixed and player ID as random effect. Plots show median (bold line) with interquartile range (box) and 1.5 fold interquartile range (whiskers). Cal.: GrimAge2 and FitAge predictions were calibrated to the actual age range of players.

Conversely, the biological ages of the supporting staff who were not exposed to the same intensity of physical stress remained constant before and after the game (Supplementary Figure 1,2). Both biological age predictors, DNAmGrimAge2 and DNAmFitAge, showed a transient change in athletes during competition, as their biological age returned to comparable values after the rest phase. Of note, also significant changes in DNAm-based Skin & Blood Clock, a well-established predictor for chronological age, were observed (Figure 2C).

A more detailed investigation of player subgroups based on their assigned positions during games revealed that midfielders experienced the most significant rejuvenation effect (Supplementary Figure 2A,C). The median DNAmGrimAge2 cal. of midfielders decreased by approximately 17.8 years (p = 0.00414) from before to after the game. This change was more pronounced compared to the moderate reductions observed in athletes playing forward (−11.3 years, p = 0.00791) and defenders (−5.3 years, p = 0.0744) (Supplementary Figure 2A). Among the supporting staff members, physicians and physiotherapists exhibited a similar trend to athletes in terms of biological age, despite no significant overall change.

### 2.2 Exercise-induced epigenetic changes reflect immunological events

Next, we aimed to investigate the epigenetic events underlying the substantial changes in DNAmGrimAge2 cal. following intense athletic workload. To this end, the influence of various plasma protein surrogate markers on changes in epigenetic age predictions induced by physical activity were examined (Figure 3). DNAmGrimAge2 consists of a group of nine DNAm-based surrogates, which were trained to predict plasma protein levels. Among the six DNAmGrimAge2 covariates significantly associated with physical activity (p = 6.13e-05), four correspond to proteins involved in inflammatory processes. Notably, a significant decrease (p = 6.78e-07) in DNAm-derived estimates of C-reactive protein (CRP) levels, but an increase in interleukin-6 (IL-6) (p = 4.81e-06) was observed post-competition among athletes, followed by a return to baseline levels after a period of rest (Figure 3A,B). In contrast, among supporting staff members, no significant alterations were observed in CRP or IL-6 levels (Supplementary Figure 1D,E). These findings suggest that activity-induced modifications in epigenetic age predictions may mirror immunologic events associated with physical exertion. The elevation of inflammatory markers in athletes post-competition could be attributed to the physiological stress and immune response triggered by high-intensity exercise. This is also corroborated by changes on cellular level. DNAm-based estimation of immune cell composition in saliva samples indicates a significant decrease in CD4 T-cells (−68%, p = 9.74e-06), whereas granulocytes increased (+44%, p = 6.78e-06) comparing before to after game samples collected from athletes (Figure 3C,D).

**Figure 3:**
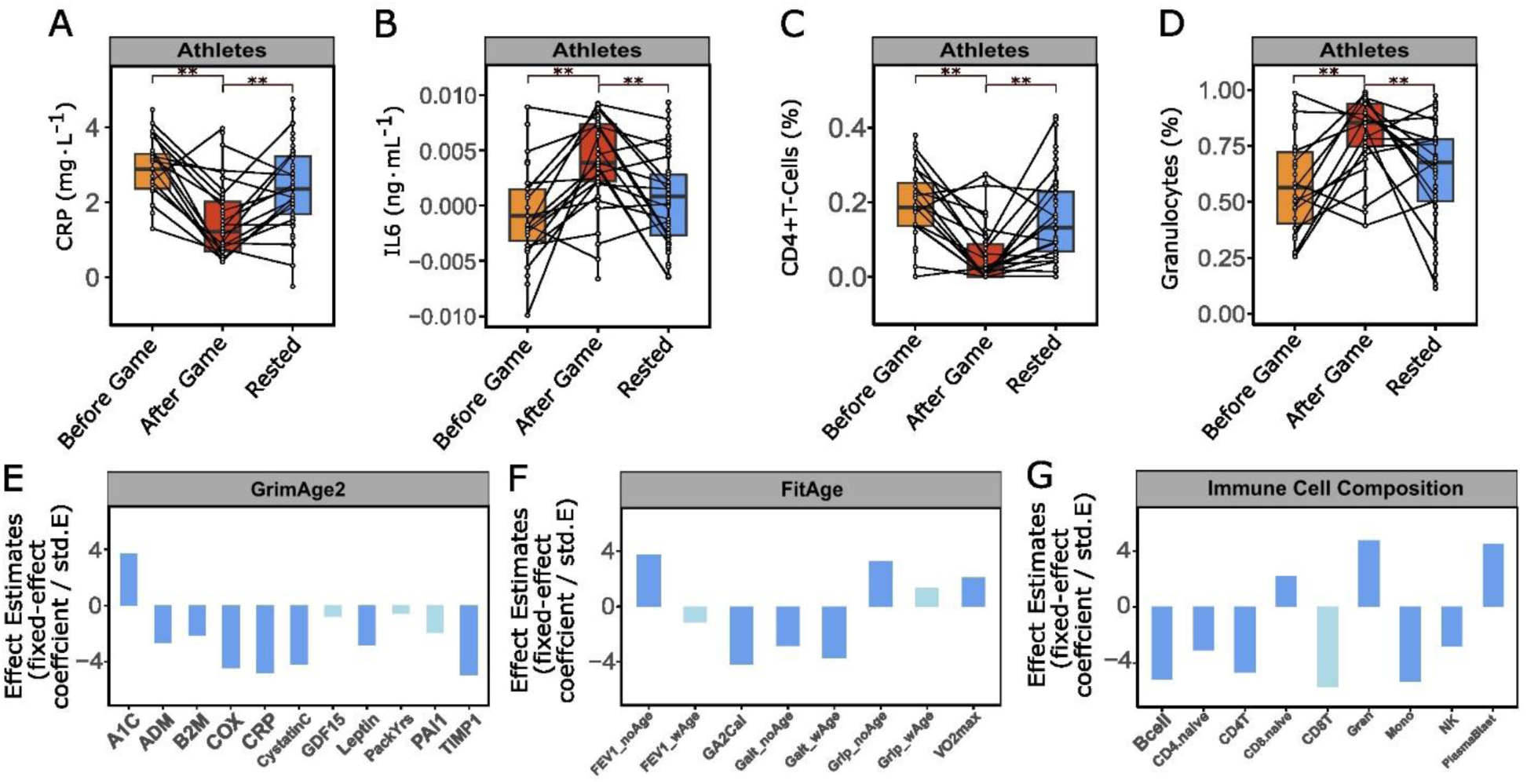
Activity-induced changes in epigenetic age predictions reflect immunologic events. (A-D) Boxplots illustrate changes of DNAm-derived surrogate estimates for blood protein levels and relative immune cell abundance before and after physical activity (before/after game) and after recovery (rested) for inflammation markers **(A)** CRP (before vs. after game p = 6.78e-07; after game vs. rested p = 0.00042) and **(B)** IL-6 (before vs. after game p = 4.81e-06; after game vs. rested p = 0.00162) as well as immune cell activity for **(C)** active CD4 T-Cells (before vs. after game p = 9.74e-06; after game vs. rested p = 0.00116) and **(D)** Granulocytes (before vs. after game p = 6.78e-06; after game vs. rested p = 0.000648), in either athletes. Each dot represents one sample from one proband, samples from the same proband are connected by line across physical activity groups, significant changes (p-values) were tested using a linear mixed effect model with chronological age, timepoint (before, after game or rested) and batch number as fixed and player ID as random effect. **(A-D)** Significance levels are indicated by * (p <= 0.05), ** (p <= 0.01) and NS. (p > 0.05). **(E-G)** Importance analysis of various clock components including DNAm-based plasma protein surrogate factor and blood immune cell composition estimates on determination of **(E)** DNAmGrimAge2, **(F)** DNAmFitAge and **(G)** DNAm based Immune Cell Composition in athletes after high intensity exercise (before vs after game). Barplot shows standardized effect estimates of various clock components (fixed effects): fixed-effect coefficients (ß) divided by their respective standard errors (std.E) from a linear regression model fitting of the marker at before against after game timepoints. DNAm-based predictor for blood levels of ADM: for adrenomedullin (ADM) (corr: −15.51, std.E: 5.84, p = 0.00924); B2M: DNAm of the beta-2 microglobulin (B2M) gene (corr: −88973.44, std.E: 41988.41, p = 0.0365); CystatinC: DNAm of the cystatin C gene (corr: −54129.71, std.E: 12900.31, p = 5.79E-05); GDF15: DNAm of the growth differentiation factor 15 (GDF15) gene (corr: −53.36, std.E: 68.41, p = 0.437); Leptin: DNAm of the leptin gene (corr: −4484.99, std.E: 1598.36, p = 0.006); PackYrs: DNAm-based estimate of smoked cigarette packs per year (corr: −0.081, std.E: 0.14, p = 0.956); PAI1: DNAm of the plasminogen activator inhibitor-1 gene (corr: −1464.13, std.E: 760.98, p = 0.0671); TIMP1: DNAm of the tissue inhibitor of metalloproteinases 1 gene (corr: −1238.74, std.E: 250.35, p = 2.96E-06); COX: A composite clinical marker (corr: −1.07, std.E: 0.24, p = 2.13E-05); A1C: DNAm-based logarithmic transformation of glycated hemoglobin (HbA1c) (corr: 0.041, std.E: 0.011, p = 0.000345); CRP: DNAm-based logarithmic transformation of C-reactive protein (corr: −1.24, std.E: 0.26, p = 4.81E-06); GA2Cal: Calibrated DNAm GrimAge, version 2, a predictor of biological age (corr: −7.07, std.E: 1.69, p = 6.13E-05); VO2max: DNAm-based estimate of maximal oxygen uptake (corr: 1.18, std.E: 0.57, p = 0.0413); Gait_noAge: DNAm-based estimate of gait, excluding age as a factor (corr: −0.085, std.E: 0.030, p = 0.00593); Grip_noAge: DNAm-based estimate of grip strength, excluding age as a factor (corr: 3.07, std.E: 0.95, p = 0.00167); FEV1_noAge: DNAm-based estimate of forced expiratory volume in one second (FEV1), excluding age as a factor (corr: 0.35, std.E: 0.095, p = 0.000307); Gait_wAge: DNAm- based estimate of gait, including age effects (corr: −0.087, std.E: 0.023, p = 0.000317); Grip_wAge: DNAm-based estimate of grip strength, including age effects (corr: 0.41, std.E: 0.30, p = 0.18); FEV1_wAge: DNAm-based estimate of FEV1, including age effects (corr: −0.051, std.E: 0.046, p = 0.268); Bcell: DNAm-based proportion estimate of B cells (corr: −0.043, std.E: 0.0083, p = 1.24E-06); CD4.naive: DNAm-based estimate proportion of naive CD4+ T cells (corr: −94.36, std.E: 30.69, p = 0.00268); CD4T: DNAm-based proportion of CD4+ T cells (corr: −0.11, std.E: 0.024, p = 9.74E-06); CD8.naive: DNAm-based proportion estimate of naive CD8+ T cells (corr: 25.43, std.E: 11.73, p = 0.0325); CD8T: DNAm-based proportion estimate of CD8+ T cells (corr: −9.51E-18, std.E: 1.68E-18, p = 1); Gran: DNAm-based proportion estimate of granulocytes (corr: 0.23, std.E: 0.049, p = 6.78E-06); Mono: DNAm-based proportion of monocytes (corr: −0.084, std.E: 0.016, p = 6.25E-07); NK: DNAm-based proportion estimate of natural killer cells (corr: −0.0070, std.E: 0.0025, p = 0.00613); PlasmaBlast: DNAm-based proportion estimate of plasmablasts (corr: 0.50, std.E: 0.11, p = 1.98E-05). Significance levels are indicated by dark blue (p <= 0.05) and light blue (p > 0.05).

Our data suggest that epigenetic age predictions based on DNA methylation events can capture immunologic changes associated with physical activity and may have implications for comprehending the effects of strenuous exercise.

### 2.3 Dynamics of short-term effects of physical exertion on epigenetic age

The reported effects of strenuous physical activity on DNA methylation-based age predictors and immune-related factors among athletes were derived from pooled samples collected throughout the season at varying intervals between high-intensity matches, training sessions, and rest phases (Figure 1,2). For a detailed examination of the short-term dynamics of epigenetic changes, we subsequently analyzed samples obtained during a 48-hour period encompassing a mid-season game, which reflected a short sequence involving low (sample 3) to high load (sample 4) and a return to resting state (sample 5) (Figure 1). Understanding the temporal dynamic of effects can contribute to developing personalized strategies for optimizing athletic performance and mitigating potential health risks associated with intensive exercise. Our data demonstrate marked changes in epigenetic age predictors DNAmGrimAge2 cal. (−31%, p = 0.0027) and DNAmFitAge cal. (−18%, p = 0.0018) immediately following intensive physical activity (i.e., straight after the game) compared with 24 hours before the game. This observation highlights the immediate impact of physical exertion on biological aging predictors (Figure 4A,B). Plasma protein surrogate factors also showed significant variation in inflammatory responses after strenuous exercise (DNAmCRP: −50%, p = 0.0018 and DNAmIL-6: +684%, p = 1.06e-05) within 24 hours (Figure 4D,E). DNAm-based immune cell type estimates exhibit comparable transient alterations in response to physical exercise (Figure 4F,G). The observed changes were of a temporary nature, with values returning to baseline levels 24 hours after the match. These values were similar to those measured 24 hours before the game.

**Figure 4:**
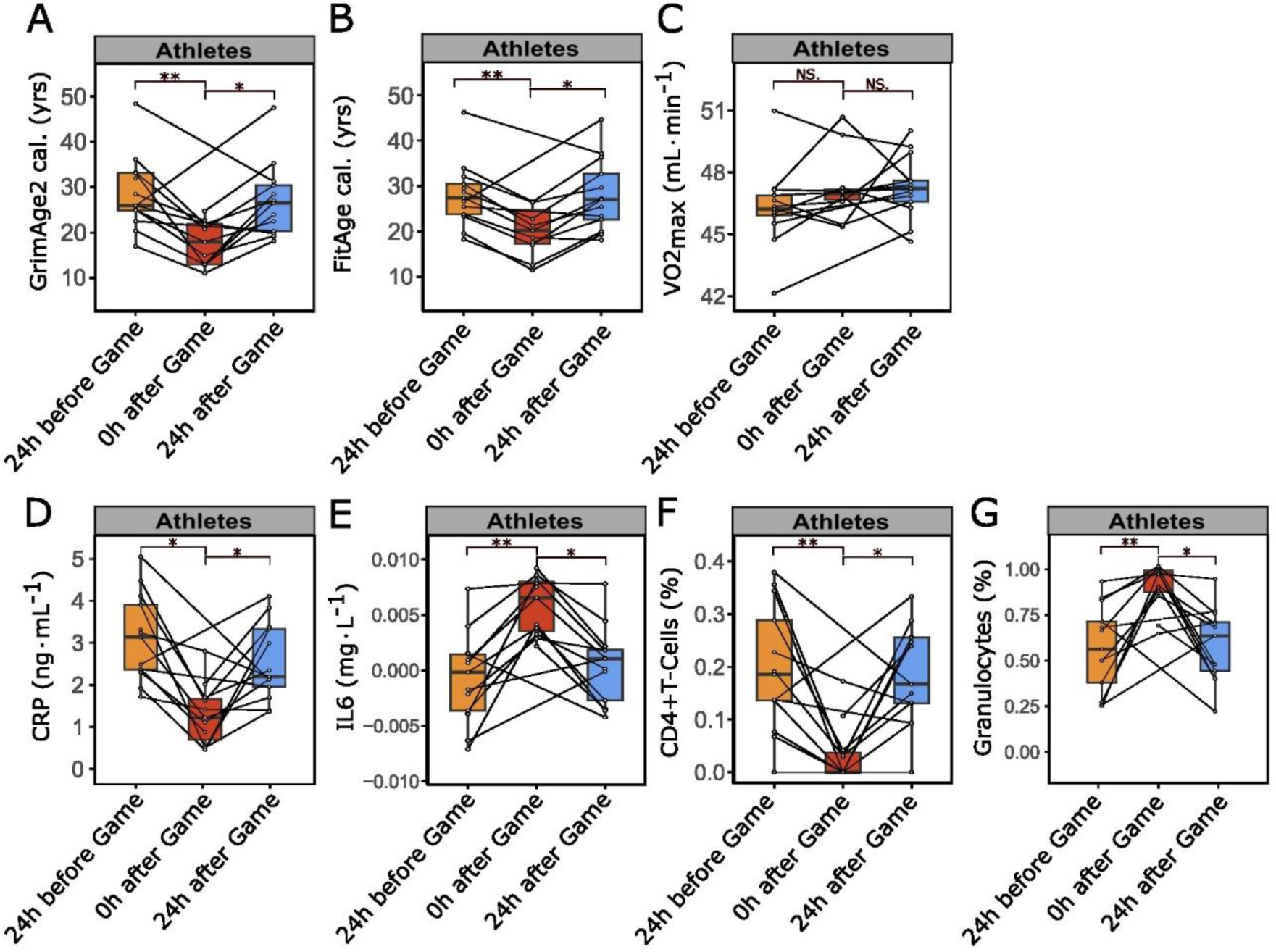
Instantaneous effects of high physical load on DNAm-based age predictors and DNAm-derived estimators of immunological factors. (A-B) Epigenetic profiles (DNAm) of saliva samples from athletes, collected during mid-season game (samples 3,4 and 5) 24 hrs before (24 hrs before game) or immediately after intensive physical activity (straight after game), were analyzed in addition to samples taken 24 hrs post high physical strain (24 hrs after game). The data was used to estimate DNAm-based biological age **(A)** GrimAge2 (24hrs before vs. straight after game p = 0.0027; straight after vs. 24 hrs after game p = 0.0166) and **(B)** FitAge (24hrs before vs. straight after game p = 0.00177; straight after vs. 24 hrs after game p = 0.0327). Analysis of DNAm-based endurance estimator **(C)** VO2max (24hrs before vs. straight after game p = 0.480; straight after vs. 24 hrs after game p = 0.274), plasma protein surrogate factors **(D)** CRP (24hrs before vs. straight after game p = 0.00177; straight after vs. 24 hrs after game p = 0.0327) and **(E)** IL-6 (24hrs before vs. straight after game p = 1.06e-05; straight after vs. 24 hrs after game p = 0.0120) and immune cell type estimates for **(F)** CD4+T-Cells (24hrs before vs. straight after game p = 0.00071; straight after vs. 24 hrs after game p = 0.0160) and **(G)** Granulocytes (24hrs before vs. straight after game p = 0.00068; straight after vs. 24 hrs after game p = 0.0118). Each dot represents one sample from one proband, samples from the same proband are connected by line across physical activity groups, significant changes (p- values) were tested using a linear mixed effect model with chronological age, timepoint (24 hrs before, straight after or 24 hrs after game) and batch number as fixed and player id as random effect. Significance levels are indicated by * (p <= 0.05), ** (p <= 0.01) and NS. (p > 0.05).

The control group of supporting staff had no significant changes in DNAmGrimAge2 cal., DNAmFitAge cal., DNAm-based immune cell type alterations and plasma protein surrogate factors (Supplementary Figure 3,4B-D). Furthermore, we conducted a comparative analysis of samples collected at the conclusion of the season against those obtained at the beginning of the season (Supplementary Figure 5). Following a 12- month period, athletes exhibited a modest trend in reduced DNAmGrimAge2 cal. (+8.8%, p=0.61), DNAmFitAge cal. (+2.7%, p=0.75) and CRP level (+16%, p=0.32) (Supplementary Figure 5A-C).

### 2.4 Co-occurrence of changes in DNA methylation and injury incidents in athletes

The analysis of closely consecutive load changes during a midseason game (samples 3-5, Figure 1) suggested an interplay between physical activity, epigenetic alterations, and immune reactions, implying a dynamic relationship among these factors. During the investigation of the changes in DNA methylation age predictors, specifically DNAmGrimAge2 cal. and DNAmFitAge cal., it was noted that the dynamic alterations typically followed a reduction from pre-game (baseline) levels to post-game levels, succeeded by a recovery back to baseline levels within the rest phase. Nonetheless, when comparing DNAmGrimAge2 cal. in players before the game (24 hrs prior, sample 3) to their rested state (24 hrs post-game, sample 5), a variable trend was observed for certain players (Supplementary Figure 6). Most players exhibited a moderate decrease in DNAmGrimAge2 cal., while a few players showed the opposite trend. Based on the fact that changes in DNAmGrimAge2 cal. were shown to correlate with immunologic events (Figure 3), which can be induced by major but also microtraumas (Gebhard et al. 2000, Schild et al. 2016), we took episodes of injuries affecting players close in time to the midseason match (Table 2) into consideration to delineate a possible source of the observed variation.

**Table 2:** Players with episodes of injury. . Player column shows anonymized participant IDs; Type of injury column shows injuries sustained during professional soccer season 2021/22 and 2022/23 matches or training sessions; Timepoint column shows timepoints of injury.

| Player | Type of injury | Timepoint |
| --- | --- | --- |
| 2 | Musculoskeletal Trauma | Sample 1, 7, 8 |
| 27 | Musculoskeletal Trauma | Samples 1-3 |
| 31 | Musculoskeletal Trauma | Sample 4 |
| 33 | Musculoskeletal Trauma | Samples 6-7 |

To this end, blood samples collected during a midseason match (Figure 1, samples 3-5) were analyzed. In specific levels of creatine kinase (CK), a marker of inflammation and tissue damage (Romagnoli et al. 2016), were measured. Additionally, DNA methylation status of saliva samples was assessed 24 hours before and after the game to evaluate changes induced by physical activity. Based on their injury history (Table 2), players were divided into two groups: those affected by musculoskeletal trauma injuries and those without. In the injury group, CK levels significantly (p = 0.013) increased on average 1.9-fold in all athletes (Figure 5A). In contrast, the non-injury group exhibited no discernible pattern in CK changes (Figure 5A). Biological age predictor DNAmFitAge cal. showed a trend of overall increase in the injury group (p = 0.257) and a decrease in the non-injury group (p = 0.174) (see Figure 5B). The only player (ID: 27) with a history of injury who didn’t show an increase in DNAmFitAge (Figure 5B, injury group panel) had suffered his injury at the beginning of the season (samples 1-3) and had already fully recovered at the time point of interest (midseason match, samples 3-5, Table 2). Comparing ratios of athletes with increasing or decreasing CK values and increasing or decreasing DNAmFitAge cal. within injury and non-injury groups 24h before and after the game, we found no statistical significant difference (Figure 5C,D, Supplementary Table 1). Interestingly, although the injury group included only 4 injured players, and only 3 players injured at the time point of analysis, we saw a trend towards a statistical significance comparing athlete ratios within the respective groups (p=0.238) (Figure 5D). Excluding player 27 with an injury event earlier in the season (samples 1-3), who fully recovered at the time point of interest (midseason match, samples 3-5, Table 2) decreased the p-value down to p=0.071, making it borderline significant (data not shown). This obvious trend could not be seen looking at athlete ratios with increasing or decreasing CK values in the injury and non-injury group (p=0.508) (Figure 5C, Supplementary Table 1).

**Figure 5:**
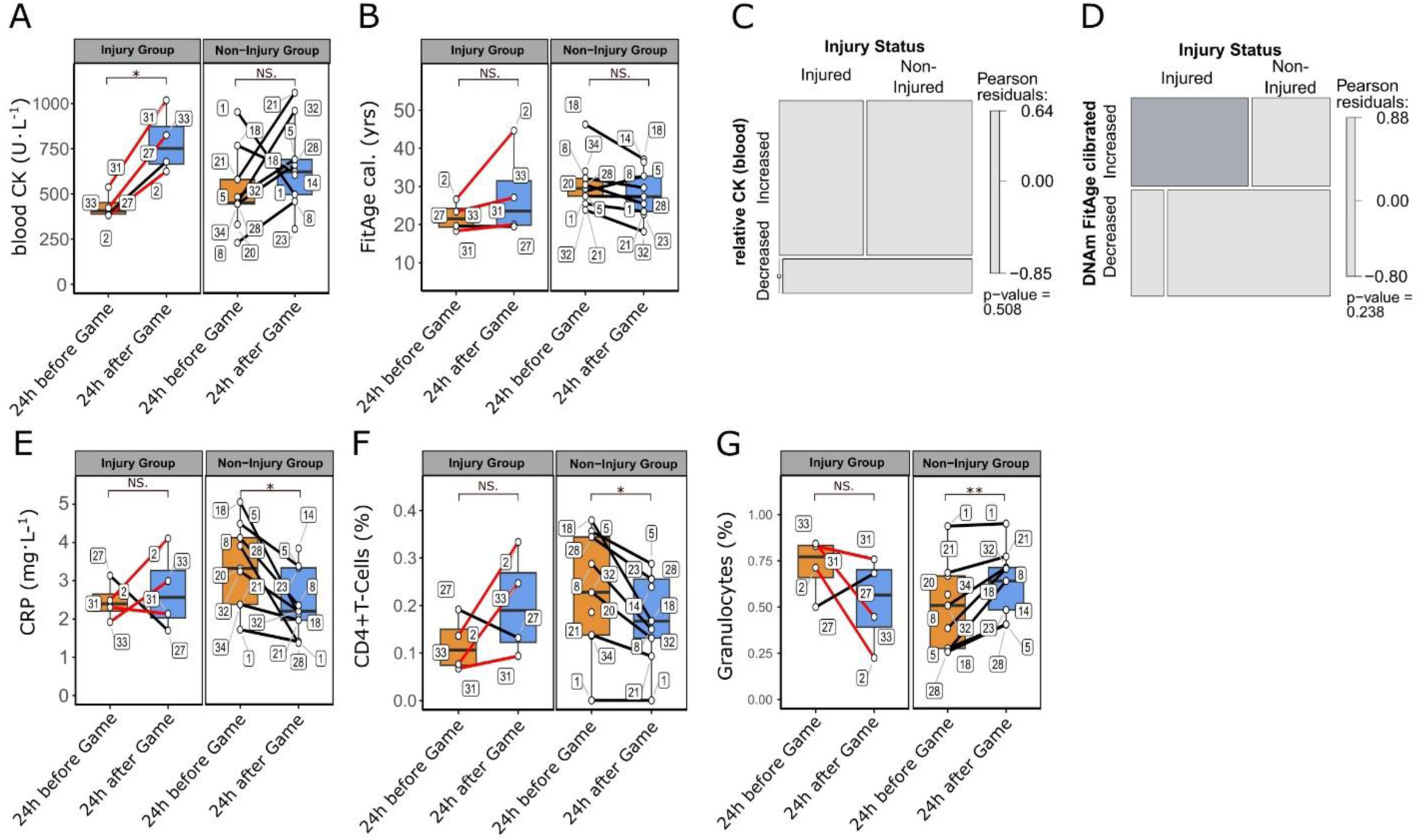
Differential patterns of biological age predictor FitAge in athletes are associated with events of injuries. In the course of a midseason match (sample 4 and 6) blood levels of inflammatory and tissue damage marker CK were determined and DNAm status of saliva samples assessed 24 hrs before and after the game. Individual players indicated by number labels in plots were split into two groups depending on injury events (red lines: players with history of injury during or after midseason match, black lines: players without injury or injury before midseason match) with temporal proximity to the match day (see Table 2). Boxplots show **(A)** CK levels (Injury Group: 24 hrs before vs. 24 hrs after game p = 0.013; Non-Injury Group: Injury Group: 24 hrs before vs. 24 hrs after game p = 0.374) and **(B)** DNAm-based FitAge cal. prediction in athletes (Injury Group: 24 hrs before vs. 24 hrs after game p = 0.257; Non-Injury Group: Injury Group: 24 hrs before vs. 24 hrs after game p = 0.174). **(C-D)** Mosaic plots illustrate ratios of athletes in injury and non-injury groups as well as the physical activity induced trend in **(C)** CK and **(D)** FitAge calibrated. Pearson residuals compare observed vs expected number of players in subgroups and highlight significant increases (blue) and decreases (red) thereof. **(E-G)** Saliva sample derived DNAm-based estimators of **(E)** inflammatory marker CRP (Injury Group: 24 hrs before vs. 24 hrs after game p = 0.657; Non-Injury Group: Injury Group: 24 hrs before vs. 24 hrs after game p = 0.020), **(F)** CD4+T-cells (Injury Group: 24 hrs before vs. 24 hrs after game p = 0.142; Non-Injury Group: Injury Group: 24 hrs before vs. 24 hrs after game p = 0.013) and **(G)** Granulocytes (Injury Group: 24 hrs before vs. 24 hrs after game p = 0.175; Non-Injury Group: Injury Group: 24 hrs before vs. 24 hrs after game p = 0.010). Each dot represents one sample from one proband, samples from the same proband are connected by line across physical activity groups, significant changes (p-values) were tested using a linear mixed effect model with chronological age, timepoint (24 hrs before or 24 hrs after game) and batch number as fixed and player id as random effect and injury status instead of timepoints for intergroup comparison. Significance levels are indicated by * (p <= 0.05), ** (p <= 0.01) and NS. (p > 0.05).

DNAm-based plasma protein surrogate factors (Figure 5E, Supplementary Figure 7) as well as immune cell composition type estimates (Figure 5F,G) resemble injury-related trends in DNAmFitAge cal. and DNAmGrimAge2 cal. patterns. This demonstrates the ability of epigenetic clocks as ensemble biomarkers to integrate the various single DNAm predictors in reflecting immunologic events related to injury.

In summary, the results highlight the potential of using DNAm-based biological age and inflammatory markers in predicting injury risk in athletes when exposed to high intensity physical load, which could lead to the development of fatigue monitoring and personalized injury prevention strategies.

## 3 Discussion

Our explorative study aimed at investigating the influence of physical and environmental stressors on athletes’ epigenetic aging determined by assessing DNAm of saliva samples collected from professional soccer players throughout a season. Instead of analyzing individual markers, we used the methylome to determine epigenetic clocks, which are determined from a combination of several factors. Indeed, our study shows that both, DNAmGrimAge2 and DNAmFitAge were able to capture transient short-term changes in athletes within 48 hours of a midseason competition as well as long-term effects throughout the entire season.

Epigenetic modifiers can affect responses to exercise training and might influence the predisposition to injury or disease (Tarnowski et al. 2022). Especially epigenetic regulation of genes highly involved in sports performances and exercise physiology, such as myocyte enhancer factor 2 (Potthoff et al. 2007) or slow-twitching type I myosin heavy chain (Pandorf et al. 2009, Barres et al. 2012) came into the focus of sports-related research. Epigenetic factors, like DNA methylation, are specific regulators of gene expression that constitute key links between the genotype, phenotypic plasticity, and environment. In the context of athlete performances, a broad spectrum of environmental factors—the physical activity itself, nutrition, emotional challenges, and pre-existing epigenetic signatures—can determine how an individual reacts to certain stressors (Ecker et al. 2018, Nagata et al. 2024).

Furthermore, physical exercise induces adaptations in the immune system and metabolic changes, with upregulation of certain enzymatic and protein factors. If physical exercise is intense or extreme, the immune response is similar to that caused by other stressors, which might be not beneficial for the athlete. Blood levels of proteins among other biomarkers that are part of the acute inflammatory response, including myoglobin, troponin, creatine kinase, lactate dehydrogenase, and C-reactive protein, are considerably increased after endurance or highly demanding sport (Bernat-Adell et al. 2021). DNAmGrimAge2 and DNAmFitAge consist of a group of nine DNAm-based surrogates, which were trained to predict plasma protein levels. Among the six DNAmGrimAge2 covariates significantly associated with physical activity, four, including DNAmCRP correspond to proteins involved in inflammatory processes. A multivariate linear regression analysis reported by Lu and colleagues revealed significant associations between saliva based AgeAccelGrim2 and clinically relevant measures, like high sensitivity C-reactive protein, with lower values of DNAm logCRP and AccelGrim2 representing higher levels of physical functioning (Lu et al. 2022). CRP is a hepatic acute-phase protein in tissue damage, and a marker of systemic inflammation and is associated with cardiovascular risk. Moreover, its levels have been correlated with frailty, morbidity, and mortality (Allen et al, 2015), which is also predicted and correlated with GrimAge and GrimAge2. It was found that in trained athletes, when a single exercise protocol was applied, CRP temporarily increased as the acute phase response after exercise (Kasapis et al. 2005). Interestingly our results showed an opposite overall trend analyzing short-term effects of physical intermittent strenuous bouts, leading to immediate decreased DNAm-based estimation of CRP levels, which normalize or even increase within a few hours after the game. Lower levels of CRP had been shown in longitudinal studies, in individuals exposed to higher levels of physical activity. In this context, although physical activity has been found to raise the CRP level acutely, it has been found that chronically physical activity reduces CRP levels (Kasapis et al. 2005). We could confirm the beneficial effect of long-term physical activity looking at sample 2 and 9, representing time points before and after season respectively. Following this 12-month period, athletes exhibited a modest but insignificant decrease in the median DNAmGrimAge2, FitAge and CRP level. This indicates that acute short- and intermittent long-term physical activity improves general fitness and biological age in soccer players.

Anaerobic training is typically used in a variety of sports settings and has been shown to have a significant impact on the composition of saliva (Ntovas et al. 2022). Because of the complexity of attributes required in sports such as soccer, they are considered randomized intermittent, dynamic, and skilled movement type sports. Research has consistently reported that acute bouts of endurance and resistance exercise can influence the migration of immune cells in the peripheral blood and saliva. Studies in saliva showed lymphocytosis immediately post exercise and a lymphopenia into the recovery period (Carlson et al. 2008, Carlson et al. 2017, Kraemer et al. 1996, Niemann et al. 1995, Simonson et al. 2004). It seems that among both the young and elderly, an active lifestyle is generally linked to lower numbers and proportions of memory T cells and higher numbers and proportions of naïve T cells. This is partly supported by a recent systematic review, concluding that regular structured exercise increases the number of naïve T cells in peripheral blood at rest (Campbell et al. 2018, Chao et al. 2017). Brown et al. characterized the T cell pool in young male and female adults classified as being very active well-trained soccer players and compared to young adults classified as being untrained. Untrained individuals showed the highest proportions of CD4+ and CD8+ memory T cells, and the lowest proportions of CD8+ naïve T cells, defined on the basis of CD57 and CD28 expression (Brown et al. 2014, Brown et al. 2015). In the present study DNAm-based estimation of immune cell composition in saliva samples indicates a significant increase in Granulocytes, whereas CD4 T-cells decreased comparing before to after game samples collected from athletes (Figure 3C,D). These results could be confirmed and were even more pronounced looking at short-term dynamics, comparing samples 3, 4 and 5. Long-term effects (sample 2 vs. 9) showed the same tendencies with lower estimated CD4 T-cells. It has to be mentioned that increase and/or reduction in the frequency and function of lymphocytes and other immune cells in peripheral blood and saliva in the hours following vigorous and prolonged exercise does not necessarily reflect immune activation nor suppression. Instead, increasing numbers of granulocytes together with lymphopenia of CD4+ T-cells and decreasing numbers of monocytes can represent a heightened state of immune surveillance and immune regulation driven by a preferential mobilization of cells to peripheral tissues. Furthermore, there is growing evidence from several studies in humans and rodents, indicating that exercise enhances, or at least does not suppress immune responses to *in vivo* challenge in younger and older individuals, supporting the contention that an acute bout of exercise has no detrimental immune consequences for health (Campbell et al. 2018). Therefore blood and/or saliva-based biomarkers can provide an objective individualized measure and monitoring of training load, recovery, health and immune status.

To illustrate the usefulness of epigenetic clocks, we analyzed the relationship between injury risk of individual athletes and changes in their DNAm-based clock predictions within a season. For this purpose, we considered samples collected during a midseason game. With one exception, injury events occurred within the following 1-2 weeks of training and competition after the midseason game. The only player (ID: 27) with a history of injury which did not show an increase in DNAmGrimAge2 and DNAmFitage had suffered his injury already at the beginning of the season (samples 1-3) and might have already fully recovered at the time point of measurement. Measurable parameters affected by exercise comprised changes in salivary cell numbers and cytokines and protein levels as an estimate using methylation data and protein levels in plasma. Our results show that the injury group followed an opposite trend compared to the overall trend of all analyzed samples within the season. After the game, within the early resting phase, the injury group showed increasing CRP and decreasing IL-6 levels estimated using methylation data. The non-injury group showed decreasing CRP and increasing IL-6 levels. These results point to a decreased fitness and health state within the early recovery phase of the injury group, whereas decreasing DNAmGrimAge2 and DNAmFitage within the non-injury group might indicate better and faster recovery of athletes after a game. Several proteins are affected in response to inflammatory processes, the majority showing increased levels shortly after an inflammatory reaction (Fedewa et al, 2016). IL-6 concentration increases more than other cytokines during exercise which might indicate muscle damage (Allen et al. 2015; Lightfoot and Cooper 2016). IL-6 plasma concentrations are reportedly affected by factors other than intensity, such as type and time of exercise (Gleeson et al. 2011; Baumert et al. 2016). IL-6 itself triggers the synthesis of hepatic acute-phase protein CRP in tissue damage (Velissaris et al. 2017).

CK, a protein involved in muscle metabolism, is frequently used in sports medicine as an indirect marker of muscle damage, and its concentration is generally considered a physical stress marker (Moghadam-Kia et al. 2016, Mougios et al. 2007, Marqués-Jiménez et al. 2016). CK levels have a significant variation with sex and ethnicity and also with exercise type: eccentric exercise causing more muscle damage than concentric contractions of the same vigor (Baumert et al. 2016, Moghadam-Kia et al. 2016). Analyzing CK blood serum levels of athletes within the injury and non-injury group of the early recovery phase we see elevated CK levels in both groups. Although a consistent pattern of elevated CK levels was observed in the injured group, the equivocal pattern of CK changes in the uninjured group suggests that CK was not sufficient to discriminate between injured and uninjured athletes.

## 4 Conclusion

The results of our study indicate that epigenetic changes analyzed by biological clock estimators like DNAmGrimAge2 and DNAmFitAge have potential to be utilized in prediction tools for injury predisposition in elite level soccer players or other intermittent strenuous sports. In the current study, we collected saliva samples, which are easily accessible within an athlete’s daily schedule and therefore well accepted. This is a major advantage compared to more invasive blood sampling which poses higher logistical challenges and a higher burden on athletes.

Despite numerous observational studies (changes in biomarkers in response to exercise), to the best of our knowledge, there are no large cohort studies of athletes in which exercise was controlled/adjusted by changes in biomarker levels or in which biomarkers reliably predicted an outcome such as injury. Therefore such studies are warranted to confirm our and others’ results. As a single biomarker for training management and injury prevention is rather unrealistic with regard to the complex pattern of physiological responses in different sport areas, development of a panel that includes aspects of inflammation, muscle status, and injury risk might allow for a more comprehensive picture of athletes and promote personalized training management within professional sports.

## 5 Methodology

As shown in figure 1, 10 measurements were taken over a total period of 187 days at irregular intervals, depending on the respective match days. The match events were two regular league games and one game in the international cup. The samples were taken depending on the respective match event. These were taken before the matchday, on the matchday straight within one hour after the match or one day after the matchday. These were combined into three groups. In group A (Rested), all data from the measurement times that were taken in a rested state were grouped. In group B (After Game), all data from the measurement time points corresponding to a loaded state were grouped, which took place immediately after the match or on the following day. Group C (Before Game) before a game, where there is a certain baseline load, but the players should be sufficiently rested for the match day.

Data was collected in different ways due to practicability. For group B (After Game) for the measurement times, ck measurement data were taken by samples of capillary blood only one day after the match and saliva samples (2ml) were taken after the match using a saliva tube (Genefix, Isohelix). Group A (Rested) for the measurement time points, ck measurement data were collected before training and saliva samples (2ml) were taken before training. The exceptions were the two samples 9a and 9b, which were taken after the summer break and for which only one saliva sample (2ml) was taken.

## Study & Statistical Design

Controlled longitudinal study with 10 measurements, comparing players vs. control staff. We set epigenetic clocks as the primary endpoint, before and after each stressor and over the season as a whole. This will primarily be performed using the GrimAge (Lu 2019) and Skin & Blood Age clocks (Horvath 2018). As a secondary outcome, using available metadata, we will assess the correlation of all other key outcomes with GrimAge, including creatine kinase levels, training load within the prior 48 hours, and self-reported measures of sleep and stress.

We will use multiple epigenetic biomarkers to assess epigenetic age before and after each stressor, and the season as a whole, including GrimAge and its component risk factors, as well as DNA methylation based predictors of immune cell subsets. In order to adjust for the longitudinal nature of the data we used two approaches: a) a linear mixed model analysis where the random intercept term corresponds to trial participant and b) a linear model where the dependent variable (epigenetic age at the completion of the trial) is regressed on epigenetic age at baseline and treatment status. Bonferroni correction was used to adjust for multiple comparisons.

## Sample Collection

Fifteen players and five staff members of a professional men’s soccer team participated in the study, providing informed written consent approved by the team’s board of directors. Genetic and epigenetic analysis involved collecting mouth saliva samples at up to 10 different time points. These time points included periods after games and during rest, such as directly after game three within a week and before vacation, after one week of Christmas vacation 2021, during an extensive preseason training period, directly before a Euroleague game, directly after a Euroleague away game, the morning after that Euroleague away game, directly after a Euroleague game following a two-week antioxidant regimen, and directly after the last league game in May 2022. For saliva sample collection, participants were instructed to refrain from eating, drinking, smoking, brushing their teeth, or chewing gum for 30 minutes prior to collection. The collection involved spitting into a collection funnel attached to a saliva collection tube until the required volume was reached, excluding bubbles. The tube was then tightly capped, and the saliva was mixed with a stabilization solution by shaking the tube several times before storing at room temperature.

## DNA Methylation

DNA methylation data was generated using the Infinium MethylationEPIC BeadChip arrays (llumina, San Diego, CA), with processing of methylation arrays by Eurofins Genomics. This enables the quantitative interrogation of more than 850,000 CpG methylation loci per sample, covering all designable RefSeq genes, with CpG Island shores, non-island CpGs, CpG islands outside of coding regions and miRNA promoter regions represented. The DNA methylation array assays involved bisulfite conversion of extracted DNA using a Zymo EZ DNA methylation kit, followed by DNA amplification, labeling, and array using MethylationEPIC BeadChip array kits, and scanning of the completed Beadchip arrays for final DNA methylation readout. The technology and methods described enable ready calculation of the most informative epigenetic aging clocks developed in the Horvath lab, including GrimAge (Lu 2019), PhenoAge (Levine 2018), the original pan-tissue clock (Horvath 2013), newer measures such as the Skin & Blood Age clock (Horvath 2018). Data analysis through the Clock Foundation enables many additional quality control measures to be performed as well as calculation of DNA methylation based surrogate measures, e.g. predicting status of the immune system.

## Additional Study Monitoring

In addition to the genetic and epigenetic analyses, a range of supplementary tests and assessments were conducted to enhance the comprehensive evaluation of the participants. Capillary blood testing was performed to measure Creatine Kinase (CK), which serves as a reliable measure of muscle strain, comparable to lactate measurements obtained from spiroergometry testing. CK levels were assessed daily to enable effective monitoring of muscle stress and provide valuable insights into the participants’ training status. Daily recordings of creatine kinase levels were stored in SAP SportsOne or Excel for subsequent analysis.

## DNA Methylation

Raw methylation signal intensities were obtained using the function read.metharray.exp from the minfi v1.40.0 R package, followed by linear dye bias correction and noob background correction to address technical variation in background fluorescence signal (Aryee et al., 2014). Specifically, the β-value was computed from the intensity of methylated and unmethylated sites as the ratio of fluorescent signals. β- values were utilized in all analyses. Subsequently, we computed several human epigenetic biomarkers of aging (epigenetic clocks) and estimated cell compositions based on blood methylation data: the pan-tissue epigenetic age (referred to as DNAmHorvath) (Horvath, 2013); Hannum’s blood-based DNAm age (DNAmHannum) (Hannum et al., 2013); skin and blood clock (DNAmAgeSkinClock) (Horvath et al., 2018); DNAm of surrogate markers of telomere length (DNAmTL) (Lu, Seeboth, et al., 2019); DNAmPhenoAge (Levine et al., 2018); the mortality risk estimator DNAmGrimAge and its components (Lu, Quach, et al., 2019); DNAmGrimAge2 (Lu et al., 2022); DNAmFitAge (McGreevy et al., 2023). Data analysis conducted through the Clock Foundation enables many additional quality control measures to be performed, as well as the calculation of DNA methylation-based surrogate measures, such as predicting the status of the immune system.

As DNAmFitAge and DNAmGrimAge2 were trained using blood samples, they tend to overestimate epigenetic age when applied to saliva samples. These two clocks were calibrated using The Clock Development Foundation trained model on a reference database consisting of 1154 healthy untreated samples with known sex and age (age varies from 18 to 91) from different sample sources (blood, saliva, buccal). The trained model estimates the sample source, age, and sex of an individual to correct the overestimation trend of the original clocks.

We employed a linear mixed-effects model, utilizing Satterthwaite’s degrees of freedom method to estimate the p-values for each coefficient. Individual ID was treated as a random effect. The response variable varied depending on the analysis, encompassing epigenetic age, components of DNAmGrimAge2, or cellular estimates. Age, sample collection group (categorized as rested, straight/one day after match, and one day before match), and plate numbers served as batch effect estimators. Notably, DNAmGrimAge2 components and cellular estimates were scaled.

## Ethics Approval

The study was approved by the local Ethics Committee of the Faculty of Psychology and Sport Science, Goethe-University Frankfurt/Main, Germany. The investigation was conducted according to the ethical standards set by the Declaration of Helsinki (World Medical Association Declaration of Helsinki: ethical principles for medical research involving human subjects. JAMA. 2013;310(20):2191–4. doi:10.1001/jama.2013.281053 Cited in: PubMed; PMID 24141714).

## Author Contributions

F.P., B.B. and S.H. conceived and planned the study design. C.H., T.B., F.P., J.S. and W.B. conceived and planned the experiments. C.H., T.B., J.S. and F.P. carried out the sample collection.

D.L. processed the samples. B.B., J.G. and M.M. developed the algorithms and performed the computations. T.K., R.Z. and M.N. verified the analytical calculations. T.K. and R.Z. contributed to the interpretation of the results. T.K. and R.Z. took the lead in writing the manuscript. All authors provided critical feedback and helped shape the research, analysis and manuscript.

## Data Availability Statement

To protect the confidentiality of the participants, we will not able to distribute these data in the public domain.

## Permission Statement

Robert T Brooke, Thomas Kocher, Roland Zauner, Juozas Gordevicius, Milda Milčiūtė, Marc Nowakowski, Christian Haser, Thomas Blobel, Johanna Sieland, Daniel Langhoff, Winfried Banzer, Steve Horvath, and Florian Pfab have all approved the submission of this manuscript.

