## Supplementary for "Epigenetic Age Monitoring in Professional Soccer Players for Tracking Recovery and the Effects of Strenuous Exercise"

### Supplementary Information

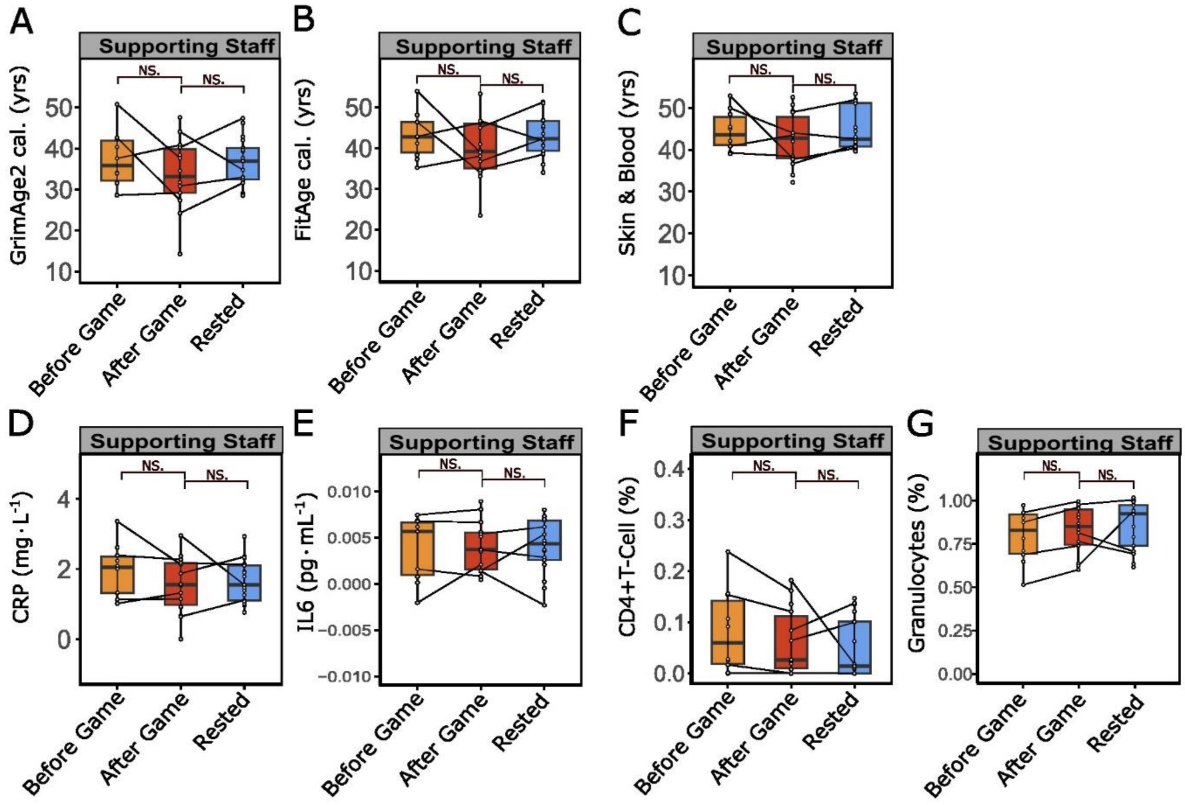


**Supplementary Figure 1: (A-C)** Epigenetic profiles (DNAm) of saliva samples collected from supporting staff members at resting states (before game, rested) or immediately after intensive physical activity (after game) were used to estimate DNAm-based biological aging clocks. **(A)** GrimAge2 cal. (before vs. after game p = 0.265; after game vs. rested p = 0.298), **(B)** FitAge cal. (before vs. after game p = 0.242; after game to rested p = 0.328) and (C) chronological age predictor Skin & Blood Clock (before vs. after game p = 0.480; after game vs. rested p = 0.299). **(D-G)** Boxplots illustrate changes of DNAm-derived surrogate blood protein levels before and after physical activity (before/after game) and after recovery (after rest) for inflammation markers **(D)** CRP (before vs. after game p = 0.227; after game vs. rested p = 0.791) and **(E)** IL-6 (before vs. after game p = 0.739; after game vs. rested p = 0.5169) as well as immune cell activity for **(F)** active CD4 T-Cells (before vs. after game p = 0.280; after game vs. rested p = 0.755) and **(G)** Granulocytes (before vs. after game p = 0.358; after game vs. rested p = 0.709). **(A-G)** Each dot represents one sample from one proband, samples from the same proband are connected by line across physical activity groups, significant changes (p-values) were tested using a linear mixed effect model with chronological age, timepoint (before game, after game or rested) and batch number as fixed and player id as random effect. Plots show median (bold line) with interquartile range (box) and 1.5 fold interquartile range (whiskers). Significance levels are indicated by * (p <= 0.05), ** (p <= 0.01) and NS. (p > 0.05).

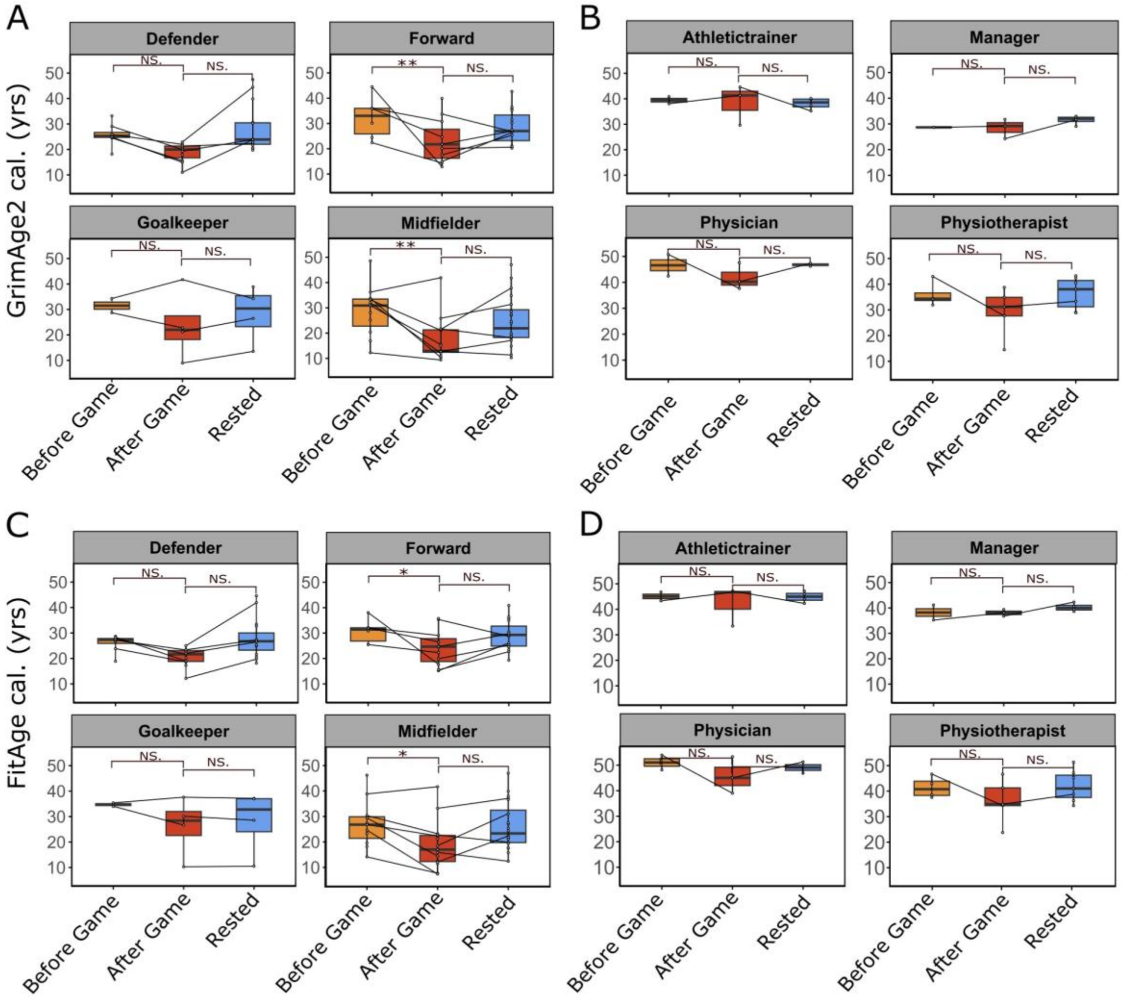


**Supplementary Figure 2: (A-D)** Epigenetic profiles of saliva samples collected from athletes at resting states (before game, rested) or immediately after intensive physical activity (after game) were used to estimate DNAm-based biological aging clocks. **(A)** GrimAge2 cal. for athletes playing different positions (before vs. after game: Defender, p = 0.074 | Forward, p = 0.00791 | Goalkeeper, p = 0.644 | Midfielder, p = 0.00414 ; after game vs. rested: Defender, p = 0.585 | Forward, p = 0.130 | Goalkeeper, p = 0.970| Midfielder, p = 0.0966) and **(B)** GrimAge2 cal. for Supporting Staff members (before vs. after game: Athletictrainer, p = 0.954 | Manager, p =0.9187 | Physician, p = 0.520 | Physiotherapist, p = 0.305; after game vs. rested: Athletictrainer, p = 0.489 | Manager, p = 0.312 | Physician, p = 0.837 | Physiotherapist, p = 0.987). **(C)** FitAge cal. for athletes at different positions (before vs. after game: Defender, p = 0.0461 | Forward, p = 0.0282 | Goalkeeper, p = 0.449 | Midfielder, p = 0.0110; after game vs. rested: Defender, p = 0.410 | Forward, p = 0.282 | Goalkeeper, p = 0.532| Midfielder, p = 0.483) and **(D)** FitAge cal. for Supporting Staff members (before vs. after game: Athletictrainer, p = 0.567 | Manager, p = 0.856 | Physician, p = 0.574 | Physiotherapist, p = 0.408; after game vs. rested: Athletictrainer, p = 0.644 | Manager, p = 0.336 | Physician, p = 0.575 | Physiotherapist, p = 0.990. **(A-D)** Each dot represents one sample from one proband, samples from the same proband are connected by line across physical activity groups, significant changes (p-values) were tested using a linear mixed effect model with chronological age, timepoint (before game, after game or rested) and batch number as fixed and player ID as random effect. Plots show median (bold line) with interquartile range (box) and 1.5 fold interquartile range (whiskers). Significance levels are indicated by * (p <= 0.05), ** (p <= 0.01) and NS. (p > 0.05). Cal.: GrimAge2 and FitAge predictions were calibrated to the actual age range of players.

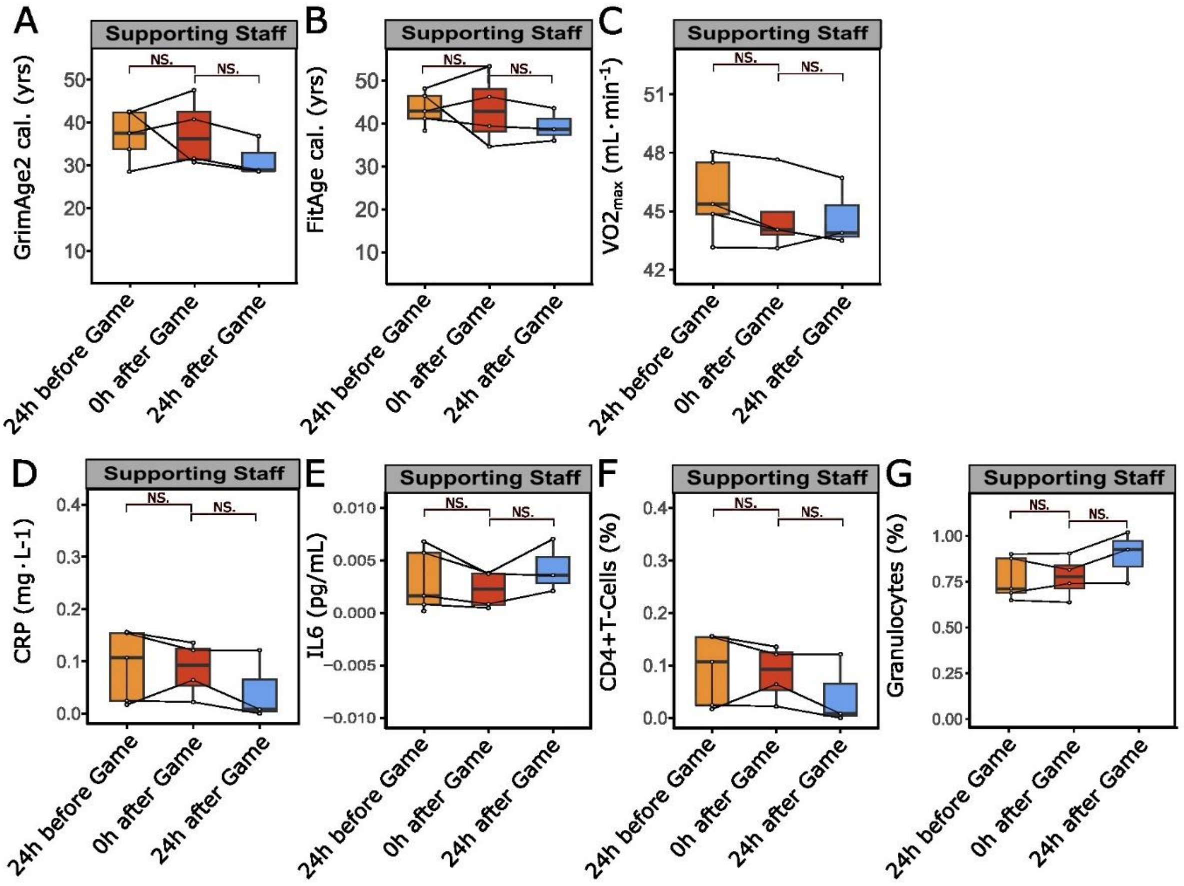


**Supplementary Figure 3: Instantaneous effects of high physical load on DNAm-derived immunological factors. (A-B)** Epigenetic profiles (DNAm) of saliva samples from supporting staff, collected as control, also during mid-season game (samples 3, 4 and 5) 24 hrs before (24 hrs before game) or immediately after the athletes underwent intensive physical activity (straight after game), were analyzed in addition to samples taken 24 hrs after the athletes high physical strain (24 hrs after game). The data was used to estimate DNAm-based biological aging clocks **(A)** GrimAge2 cal. (24hrs before vs. straight after game p = 0.944; straight after vs. 24 hrs after game p = 0.337) and **(B)** FitAge cal. (24hrs before vs. straight after game p = 0.663; straight after vs. 24 hrs after game p = 0.409). Analysis of DNAm-based endurance estimator **(C)** VO2max (24hrs before vs. straight after game p = 0.344; straight after vs. 24 hrs after game p = 0.862), plasma protein surrogate factors **(D)** CRP (24hrs before vs. straight after game p = 0.512; straight after vs. 24 hrs after game p = 0.444) and **(E)** IL-6 (24hrs before vs. straight after game p = 0.754; straight after vs. 24 hrs after game p = 0.426) and immune cell type estimates for **(F)** CD4+T-Cells (24hrs before vs. straight after game p = 0.762; straight after vs. 24 hrs after game p = 0.798) and **(G)** Granulocytes (24hrs before vs. straight after game p = 0.971; straight after vs. 24 hrs after game p = 0.623). Each dot represents one sample from one proband, samples from the same proband are connected by line across physical activity groups, significant changes (p-values) were tested using a linear mixed effect model with chronological age, timepoint (24 hrs before, straight after or 24 hrs after game) and batch number as fixed and player id as random effect. Significance levels are indicated by * (p <= 0.05), ** (p <= 0.01) and NS. (p > 0.05).

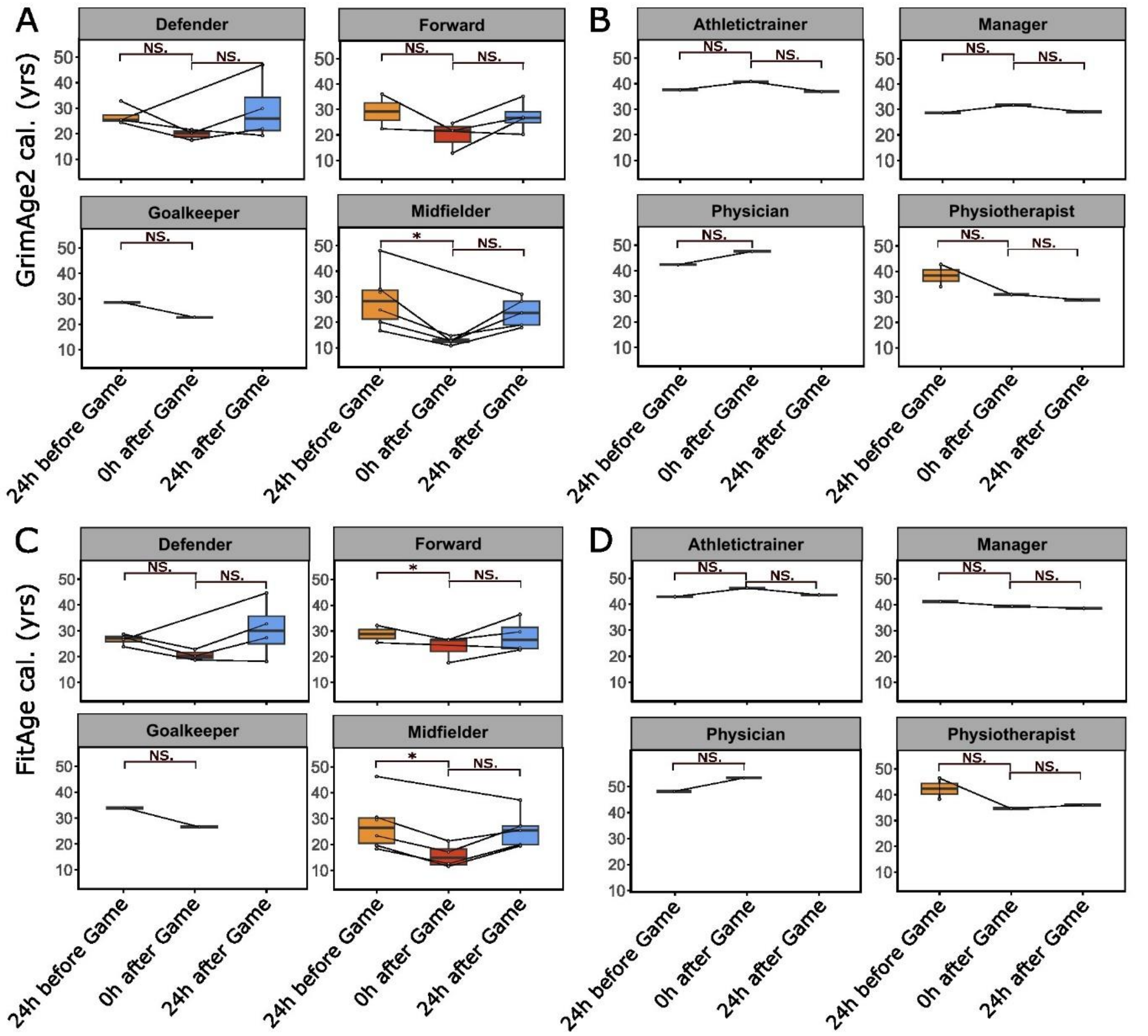


**Supplementary Figure 4: Instantaneous effects of high physical load on DNAm-based age predictors. (A-D)** Epigenetic profiles (DNAm) of saliva samples collected from athletes at resting states (24 hrs before game, 24 hrs after game) or immediately after intensive physical activity (straight after game [Str. AG]) were used to estimate DNAm-based biological aging clocks. **(A)** GrimAge2 cal. for athletes at different positions (24 hrs before vs. straight after game: Defender, p = 0.38 | Forward, p = 0.052 | Goalkeeper, p = 1.00 | Midfielder, p = 0.012; straight after game vs. 24 hrs after game: Defender, p = 0.223 | Forward, p = 0.0776 | Goalkeeper, missing data | Midfielder, p = 0.130) and **(B)** GrimAge2 cal. for Supporting Staff members (24 hrs befor vs. straight after game: Athletictrainer, p = 1.00 | Manager, p = 1.00 | Physician, missing data| Physiotherapist, p = 0.505; straight after game vs. 24hrs after game: Athletictrainer, p = 1.00 | Manager, p = 1.00 | Physician, missing data | Physiotherapist, p = 0.85; 24hrs before game vs. 24 hrs after game: Physician, p = 1.00), **(C)** FitAge cal. for athletes at different positions (24 hrs before vs. straight after game: Defender, p = 0.387 | Forward, p = 0.0282 | Goalkeeper, p = 1.00 | Midfielder, p = 0.0114; straight after game vs. 24 hrs after game: Defender, p = 0.123 | Forward, p = 0.29 | Goalkeeper, missing data | Midfielder, p = 0.0941) and **(D)** FitAge cal. for Supporting Staff members (24 hrs before vs. straight after game: Athletictrainer, p = 1.00 | Manager, p = 1.00 | Physician, missing data | Physiotherapist, p = 0.468; straight after game vs. 24 hrs after game: Athletictrainer, p = 1.00 | Manager, p = 1.00 | Physician, missing data | Physiotherapist, p = 0.891; 24hrs before game vs. 24 hrs after game: Physician, p = 1.00). **(A-D)** Each dot represents one sample from one proband, samples from the same proband are connected by line across physical activity groups, significant changes (p-values) were tested using a linear mixed effect model with chronological age, timepoint (before game, after game or rested) and batch number as fixed and player id as random effect. Plots show median (bold line) with interquartile range (box) and 1.5 fold interquartile range (whiskers). Significance levels are indicated by * (p <= 0.05), ** (p <= 0.01) and NS. (p > 0.05).

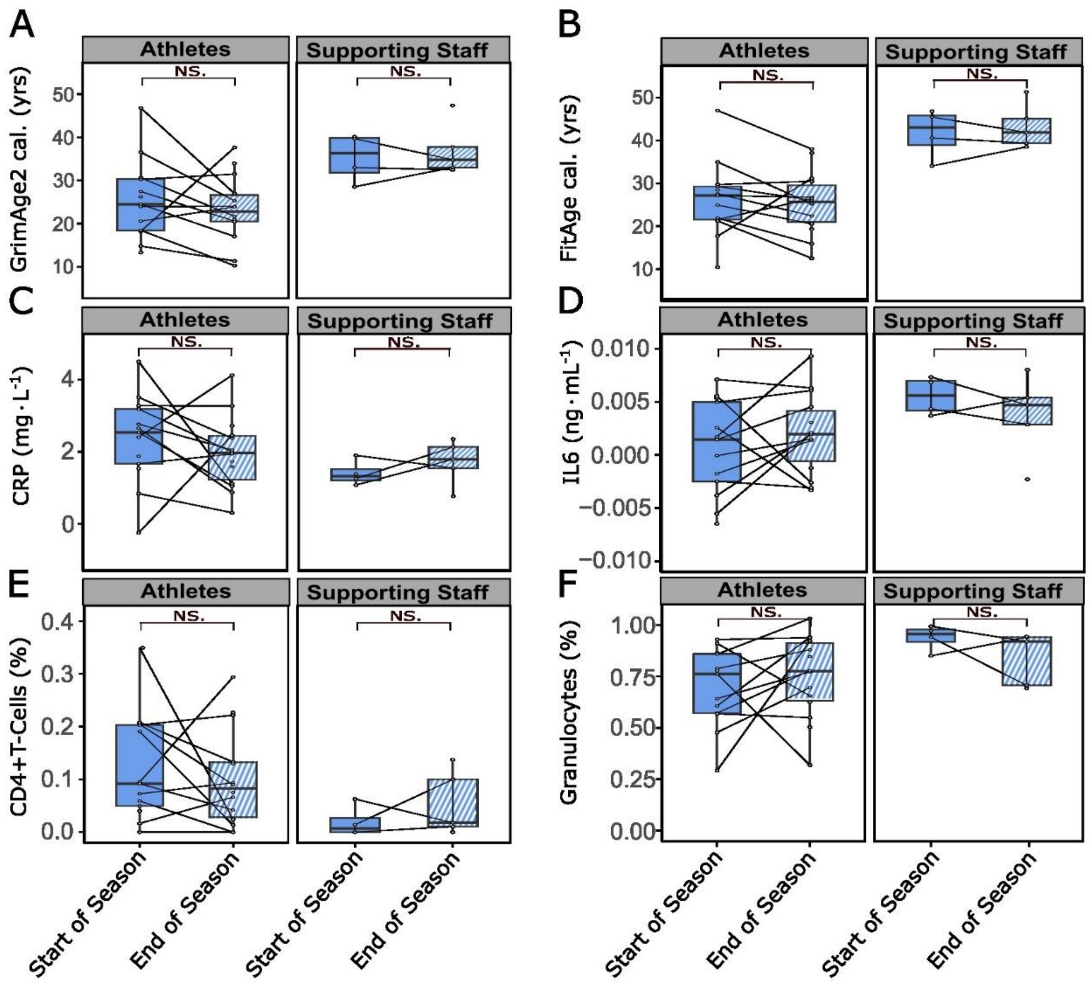


**Supplementary Figure 5: Long term effects of high physical load on DNAm-based age predictors and DNAm-derived immunological factors. (A-B)** Epigenetic profiles (DNAm) of saliva samples from athletes, collected during beginning (samples 2, blue) and end of season (sample 9, blue lined) from athlete probands and supporting staff as control. The data was used to estimate DNAm-based biological aging clocks **(A)** GrimAge2 cal. (Athletes: beginning of season vs. end of season p = 0.219; Supporting Staff: beginning of season vs. end of season p = 0.673) and **(B)** FitAge cal. (Athletes: beginning of season vs. end of season p = 0.292; Supporting Staff: beginning of season vs. end of season p = 0.673. **(C-D)** Analysis of DNAm-based plasma protein surrogate factors **(C)** CRP (Athletes: beginning of season vs. end of season p = 0.338; Supporting Staff: beginning of season vs. end of season p = 0.400) and **(D)** IL-6 (Athletes: beginning of season vs. end of season p = 0.319; Supporting Staff: beginning of season vs. end of season p = 0.592) and **(E-F)** immune cell type estimated proportion for **(E)** CD4+ T-Cells (Athletes: beginning of season vs. end of season p = 0.461; Supporting Staff: beginning of season vs. end of season p = 0.349) and **(F)** Granulocytes (Athletes: beginning of season vs. end of season p = 0.410; Supporting Staff: beginning of season vs. end of season p = 0.208). **(A-F)** Each dot represents one sample from one proband, samples from the same proband are connected by line across physical activity groups, significant changes (p-values) were tested using a linear mixed effect model with chronological age, timepoint (beginning of season, end of season) and batch number as fixed and player id as random effect. Plots show median (bold line) with interquartile range (box) and 1.5 fold interquartile range (whiskers). Significance levels are indicated by * (p <= 0.05), ** (p <= 0.01) and NS. (p > 0.05).

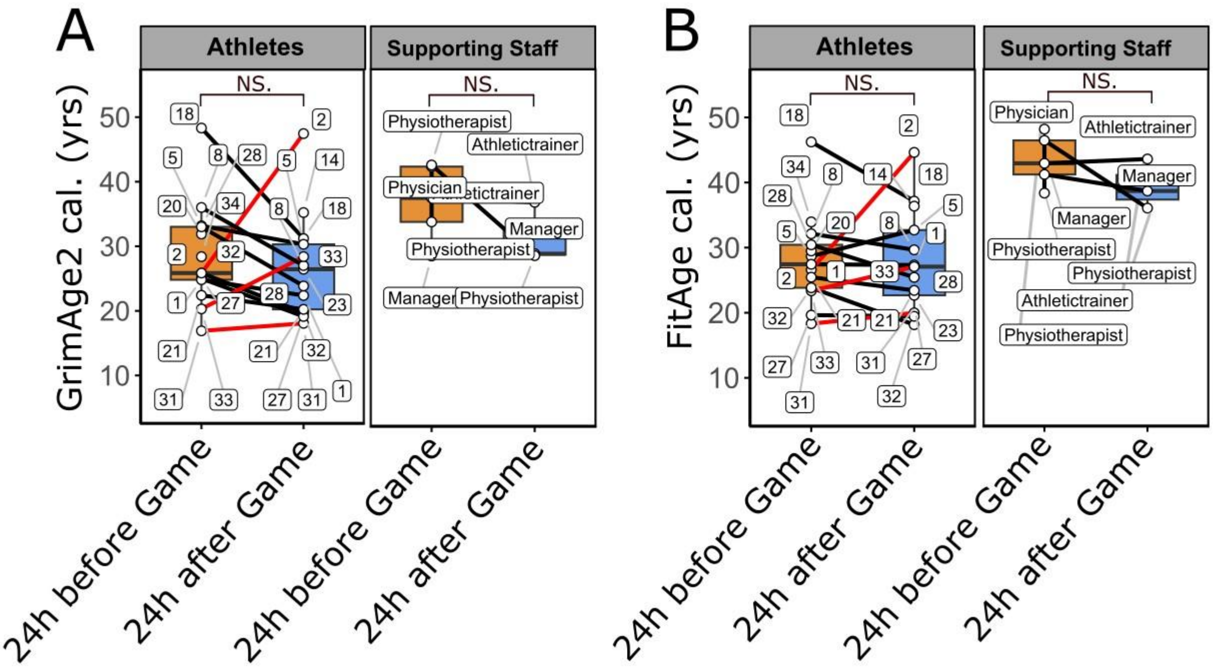


**Supplementary Figure 6: Short term effect of high physical load on DNAm-based age predictors (A-B)** Epigenetic profiles (DNAm) of saliva samples from athletes, collected during mid-season games (samples 3 and 5) 24 hrs before (before match: sample 3) and 24 h after (after match: sample 5) match day, were analyzed in addition to control samples from supporting staff members. The data was used to estimate DNAm-based biological aging clocks **(A)** GrimAge2 cal. (Athletes: 24hrs before game vs. 24hrs after game, p = 0.731; Supporting Staff: 24hrs before game vs. 24hrs after game, p = 0.212) and **(B)** FitAge cal. (Athletes: 24hrs before game vs. 24hrs after game, p = 0.77381; Supporting Staff: 24hrs before game vs. 24hrs after game, p = 0.266) for each group. Each dot represents one sample from one proband, samples from the same proband are connected by line across physical activity groups, significant changes (p-values) were tested using a linear mixed effect model with chronological age, timepoint (24hrs before game vs. 24hrs after game) and batch number as fixed and player id as random effect. Plots show median (bold line) with interquartile range (box) and 1.5 fold interquartile range (whiskers). Significance levels are indicated by * (p <= 0.05), ** (p <= 0.01) and NS. (p > 0.05).

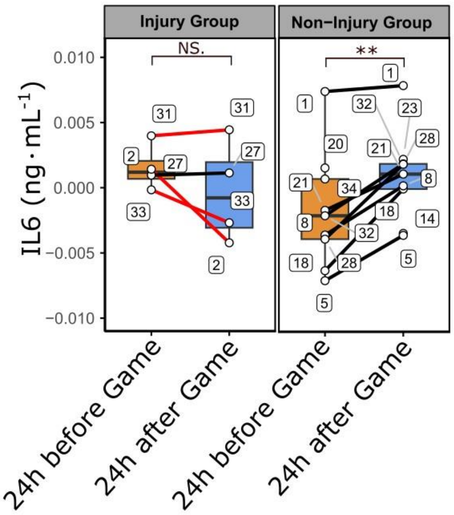


**Supplementary Figure 7: Short term effect of high physical load on DNAm-derived immunological factor IL-6.** Immunological profile of saliva samples from athletes of the injury group and non-injury group in comparison, collected during mid-season game (samples 3 and 5) 24 hrs before (before match: sample 3) and one day after (after match: Sample 5) match were analyzed. The data was used to estimate DNAm-based plasma protein surrogate factor IL6 (Injury Group: 24 hrs before game vs. 24 hrs after game, p = 0.222; Non-Injury Group: 24 hrs before game vs. 24 hrs after game, p = 0.00194). Each dot represents one sample from one proband, samples from the same proband are connected by line across physical activity groups, significant changes (p-values) were tested using a linear mixed effect model with chronological age, timepoint (beginning of season, end of season) and batch number as fixed and player id as random effect. Plots show median (bold line) with interquartile range (box) and 1.5 fold interquartile range (whiskers). Significance levels are indicated by * (p <= 0.05), ** (p <= 0.01) and NS. (p > 0.05).

**Supplementary Table 1.** Table shows direction of effect of 24 hrs before game vs. 24 hrs after game in **(A)** CK value change and **(B)** DNAmFitAge change. A table of the CK and DNamGrimAge2 status in group comparison showing the number of athlete probands suffering from an injury during season and having an increase or decrease in CK or DNAmGrimAge2 values between sample 3 and 5 with non-injury group as comparison.

A

| Status | Injury Status | |
|---|---|---|
| CK | Injured | Non-injured |
| increased | 4 | 5 |
| descreased | 0 | 2 |

B

| Status | Injury Status | |
|---|---|---|
| DNAm FitAge | Injured | Non-injured |
| increased | 3 | 1 |
| descreased | 1 | 6 |
